# Evaluation of Xpert^®^ MTB/XDR test for susceptibility testing of *Mycobacterium tuberculosis* to first and second-line drugs in Uganda

**DOI:** 10.1101/2023.04.03.23288099

**Authors:** Achilles Katamba, Willy Ssengooba, James Sserubiri, Derrick Semugenze, Kasule George William, Nyombi Abdunoor, Raymond Byaruhanga, Stavia Turyahabwe, Moses L Joloba

## Abstract

**Background:** Drug-Resistant Tuberculosis (DR-TB) is one of the key challenges toward TB control. There is an urgent need for rapid and accurate drug susceptibility tests (DST) for the most commonly used 1^st^ and 2^nd^ line TB drugs.

**Design and Methods:** In a blinded, laboratory-based cross-sectional study, we set out to validate the performance of the Xpert^®^ MTB/XDR test for DST of *M. tuberculosis*. Sputum samples or culture isolates collected between January 2020 and December 2021 from patients with rifampicin resistance –TB and/or with higher suspicion index for isoniazid (INH) resistance and/or 2^nd^ line fluoroquinolones (FQ) and injectable agents (IAs) were tested using the Xpert^®^ MTB/XDR test from 11/September 2021 to 26/May /2022. Diagnostic accuracy and factors for laboratory uptake of Xpert^®^ MTB/XDR test were compared to MGIT960 and the Hain Genotype® MTBDR*plus* and MDRsl assays (LPA) as reference DST methods.

**Results:** A total of 100 stored sputum samples were included in this study. Of the samples tested using MGIT960, 65/99 (65.6%) were resistant to INH, 5/100 (5.0%) resistant to FQ and none were resistant to IAs. The sensitivity and specificity, n (%; 95%Confidence Interval, CI) of Xpert^®^ MTB/XDR test for; INH were 58 (89.2; 79.1-95.5) and 30 (88.2; 72.5-96.6), FQ; 4 (80.0; 28.3-99.4) and 95 (100; 96.2-100), respectively. The specificity for AIs was 100 (100; 96.3-100).

Using LPA as a reference standard, a total of 52/98 (53.1%) were resistant to INH, 3/100 (3.0%) to FQ, and none to IA. The sensitivity and specificity, n (%; 95%CI) of Xpert^®^ MTB/XDR test compared to LPA for; INH was 50 (96.1; 86.7-99.5) and 34 (74.0; 58.8-85.7) and FQ 3 (100; 29.2-100) and 96 (99.0; 94.3-99.9) respectively. The specificity of IAs was 96 (100; 96.2-100).

The factors for laboratory uptake and roll-out included; no training needed for technicians with previous Xpert-ultra experience and one day for those without, recording and reporting needs were not different from those of Xpert ultra, the error rate was 4/100 (4%), no uninterpretable results reported, test turn-around-time was 1hr/45 minutes and workflow similar to that of the Xpert-ultra test.

**Conclusion:** There is high sensitivity and specificity of Xpert^®^ MTB/XDR test for isoniazid, fluoroquinolones, and Injectable agents. There are acceptable Xpert^®^ MTB/XDR test attributes for test uptake and roll-out.

## Introduction

Tuberculosis (TB), caused by *M. tuberculosis* (MTB), remains an important cause of global morbidity and mortality[1]. Moreover, the rates of drug-resistant TB (DR-TB), including TB caused by bacteria that are resistant to the most active and useful TB drugs, are increasing in many countries. The World Health Organization (WHO) TB report of 2020 reported a reduction of 5.8 million newly diagnosed with TB (−18% compared to 2019), attributed to the COVID-19 pandemic. This is likely to show that the number of people undiagnosed and untreated increased and likely DR-TB. The burden of DR-TB is also estimated to have increased between 2020 and 2021, with 450 000 (95% UI: 399 000–501 000) new cases of rifampicin-resistant TB (RR -TB) in 2021[1]. Treatment for MDR-TB is not only longer, but also more expensive, ≥US$ 1000 per person, with only a 55% success rate globally[1]. Rapid DR-TB diagnosis followed by timely initiation to appropriate treatment remains a high priority[2].

There is still a huge need to increase DR-TB diagnosis and treatment initiation. Drug susceptibility testing (DST) of MTB is increasingly important for appropriate clinical care of individual patients and successful TB control at the population level. The reference methods for MTB clinical DST are qualitative and are based upon the growth/no growth at one or two critical concentrations of antibiotic to determine the susceptibility status of an isolate. The existing standard reference methods for MTB DST are the solid agar proportions method and the liquid-based system: Mycobacterial Growth Indicator Tube (MGIT960®) system (Becton Dickinson, Sparks, Maryland). The agar proportions method relies on the use of media generally prepared within the laboratory, while the MGIT system can be purchased as a kit. There are several important problems with the existing approaches. First, these require highly specialized laboratories with advanced biosafety standards and highly skilled personnel. The procedures have technical issues as bacterial organisms tend to clump, making it difficult to prepare the testing inoculum and accurately determine colony-forming units. The drugs are usually lyophilized and require reconstitution and therefore prone to technical errors.

Therefore, a genotypic susceptibility testing methodology with the kit format and automated DNA-based technology that detect resistance-conferring mutations rather than culture-based may be advantageous. A “kit-based” approach using standardized, quality-assured antibiotics within the cartridge with automated testing will be attractive. There are alternative DNA-based methods that have been in existence for decades, but they are presently expensive, test only limited targets, are labor intensive, and require dedicated laboratory areas. These include a rapid, but less accessible molecular MDR-TB, preXDR-TB, and XDR-TB diagnostic, the line probe assay (LPA) the Genotype® MTBDR*plus* MDRsl (Hain Life Sciences, Nehren, Germany). Although LPA is rapid and offers DST results to rifampicin, isoniazid (INH), fluoroquinolones (FQs), and 2^nd^ line injectable agents (IAs), it requires extra technical skills as well as infrastructure requirements.

As part of the efforts to reduce these challenges and the diagnostic gap for MDR-TB, the WHO endorsed the use of GeneXpert MTB/RIF test (Xpert; Cepheid, Sunnyvale, CA) in 2011 as the initial diagnostic test in individuals suspected of having MDR-TB or HIV-associated TB[3]. This was followed by the recommendation of the WHO End TB strategy towards the reduction of the MDR-TB burden.

The WHO End TB strategy recommends key actions including; universal screening for drug resistance, TB treatment informed by drug resistance patterns, and the use of shorter regimens with drugs that are more effective[4]. As it is for susceptible TB, early diagnosis of MDR-TB and XDR-TB is paramount towards TB control and elimination efforts, however, this remains a challenge in most of the low and middle-income countries (LMICs)[5, 6]. The GeneXpert test has played a big part in the early diagnosis of MDR-TB in which most of the LMICs are basing MDR-TB treatment initiation on the Xpert results only i.e. patients with Xpert rifampicin resistance (Xpert-RR) detected are initiated on MDR-TB treatment.

In 2017, WHO endorsed the use of Xpert Ultra (Ultra; Cepheid, Sunnyvale, CA, USA) assay [7, 8] which is a second-generation Xpert with improved sensitivity for the diagnosis of TB as well as detection of rifampicin resistance. However, the current version of Xpert cannot determine susceptibility to INH, FQs, and 2^nd^ line IAs. Recently Cepheid has developed and validated a novel cartridge, the Xpert^®^ MTB/XDR test (launched in 2021), which is capable of determining susceptibility to these drugs. This test is currently recommended as a reflex test for high-risk patients with INH resistance as well as patients with a high risk of resistance to second-line anti-TB drugs (FQs and IAs). In 2021, WHO endorsed and recommended additional field evaluation of the Xpert^®^ MTB/XDR test before final endorsement. The current study aimed at validating the performance of the Xpert^®^ MTB/XDR test for susceptibility testing of MTB among presumptive XDR-TB patients. The test performance indicators were compared with the current standard the MGIT 960 and LPA DST methods.

## Materials and Methods

### Study design and population

This was a blinded, laboratory-based cross-sectional study to determine the performance of the Xpert^®^ MTB/XDR test in stored sputum samples collected between January 2020 and December 2021 from patients with rifampicin resistance for detection of resistance to INH and/or 2^nd^ line FQs and IAs in comparison to WHO endorsed MGIT 960 DST and/or Hain MDR*plus* and MTBDR*sl* as reference comparator. Samples included in this study were those stored and with results reported as such by the National TB Reference laboratory (NTRL) or by Mycobacteriology (BSL-3) laboratory at Makerere University. The results of the investigational Xpert^®^ MTB/XDR test were not used for clinical care. Testing with Xpert^®^ MTB/XDR test was conducted between 11/September 2021 to 26/May /2022. All operators performing investigational tests were blinded to the previous DST results of the sample. The Xpert^®^ MTB/XDR assay was performed by different operators and each operator was blinded to the other investigational test’s result. To be eligible, the stored sample had to have a corresponding MTB isolate for repeat phenotypic and genotypic DST if needed. However, efforts were made to obtain the reference standard DST results from tests previously done from the two reference laboratories. Stored sputum samples were selected from previously treated TB patients at high risk of mono isoniazid resistance and from patients diagnosed with rifampicin resistance who are at high risk of having resistance to FQs and/or IAs.

#### Laboratory procedures

All study procedures were performed according to written Standard Operating Procedures (SOPs). Well-selected stored sputum samples were checked for the availability of reference test results. Retrieved samples were processed according to the SOPs of the laboratory for the reference standard tests (MGIT960 DST and/or Hain MDR*plus* and MTBDR*sl*), in case they were missing from the testing laboratory where the sample is obtained, as well as according to the manufacturer’s instructions for the Xpert^®^ MTB/XDR test.

#### Statistical analysis

Repeat testing was done in case of discordant results defined as; a) Samples with any resistance on the Xpert^®^ MTB/XDR test but no resistance detected by any of the reference standard tests, b) Samples which have no resistance detected on Xpert^®^ MTB/XDR test but with resistance to isoniazid and/or 2^nd^ line drugs on any of the reference standard tests. For the analysis of the diagnostic accuracy analysis, the Xpert^®^ MTB/XDR test was performed on stored sputum or specimens meeting criteria for indeterminate cases after two repeats were excluded. The main analyses comparing Xpert^®^ MTB/XDR test to MGIT-960 DST and Hain MDR*plus* and MTBDR*sl* (reference standard tests) results consisted of constructing exact 95% confidence intervals around the proportions of interest: sensitivity, specificity, prediction positive and negative values. For feasibility analysis, descriptive statistics were used to calculate and compare test failure rates, the number of hours required for training, training runs required, the time required to run each test, and test indeterminate rates.

#### Ethical considerations

The study received Research Ethics committee approval from the Makerere University School of Biomedical Sciences Research Ethics Committee (SBS-2021-19) and the Uganda National Council of Science and Technology (HS1395ES). Administrative approval to use anonymized stored samples were obtained from the National TB and Leprosy programme, Ministry of Health Uganda and from the Mycobacteriology (BSL-3) laboratory, Department of Medical Microbiology, Makerere University. The study received waiver of consent since it used fully anonymized stored samples.

## Results

A total of 100 stored sputum samples were included in this study. Of the samples tested using MGIT960 phenotypic DST, 65/99 (65.6%) were resistant to isoniazid (INH), 5/100 (5.0%) resistant to fluoroquinolones (FQ) and none was resistant to the injectable agent (IA). The sensitivity and specificity, n (%; 95%Confidence Interval, CI) of the Xpert^®^ MTB/XDR test for; INH were 58 (89.2; 79.1-95.5) and 30 (88.2; 72.5-96.6), FQ 4 (80.0; 28.3-99.4) and 95 (100; 96.2-100), respectively. The specificity for AIs was 100 (100; 96.3-100), Table 1.

**Table 1:**
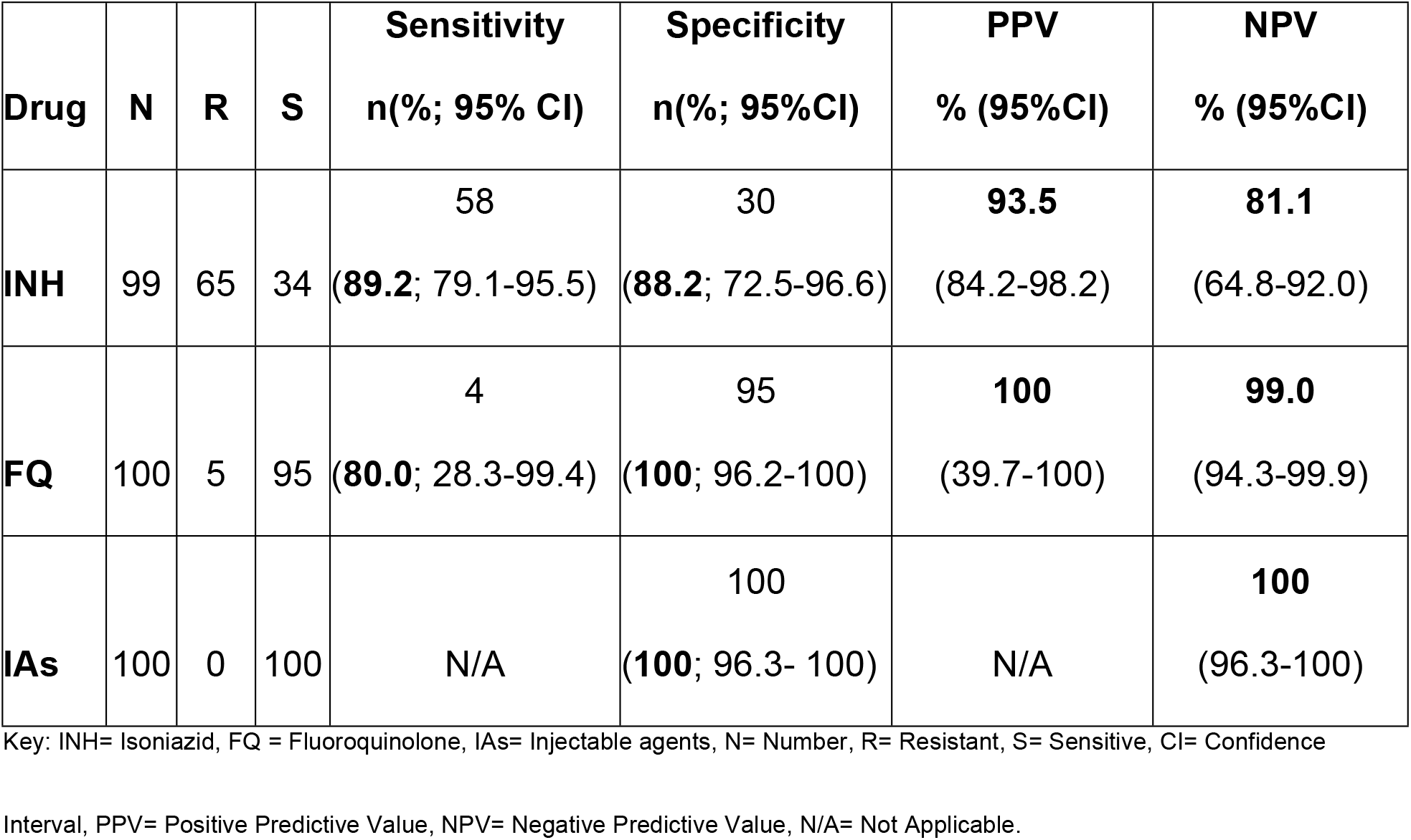
Diagnostic accuracy of Xpert^®^ MTB/XDR test using Phenotypic Drug Susceptibility test as a reference comparator.

| **Drug** | **N** | **R** | **S** | **Sensitivity**<br>**n(%; 95% CI)** | **Specificity**<br>**n(%; 95%CI)** | **PPV**<br>**% (95%CI)** | **NPV**<br>**% (95%CI)** |
| --- | --- | --- | --- | --- | --- | --- | --- |
| **INH** | 99 | 65 | 34 | 58<br>(**89.2**; 79.1-95.5) | 30<br>(**88.2**; 72.5-96.6) | **93.5**<br>(84.2-98.2) | **81.1**<br>(64.8-92.0) |
| **FQ** | 100 | 5 | 95 | 4<br>(**80.0**; 28.3-99.4) | 95<br>(**100**; 96.2-100) | **100**<br>(39.7-100) | **99.0**<br>(94.3-99.9) |
| **IAs** | 100 | 0 | 100 | N/A | 100<br>(**100**; 96.3- 100) | N/A | **100**<br>(96.3-100) |

Key: INH= Isoniazid, FQ = Fluoroquinolone, IAs= Injectable agents, N= Number, R= Resistant, S= Sensitive, CI= Confidence
Interval, PPV= Positive Predictive Value, NPV= Negative Predictive Value, N/A= Not Applicable.

**Table 2:**
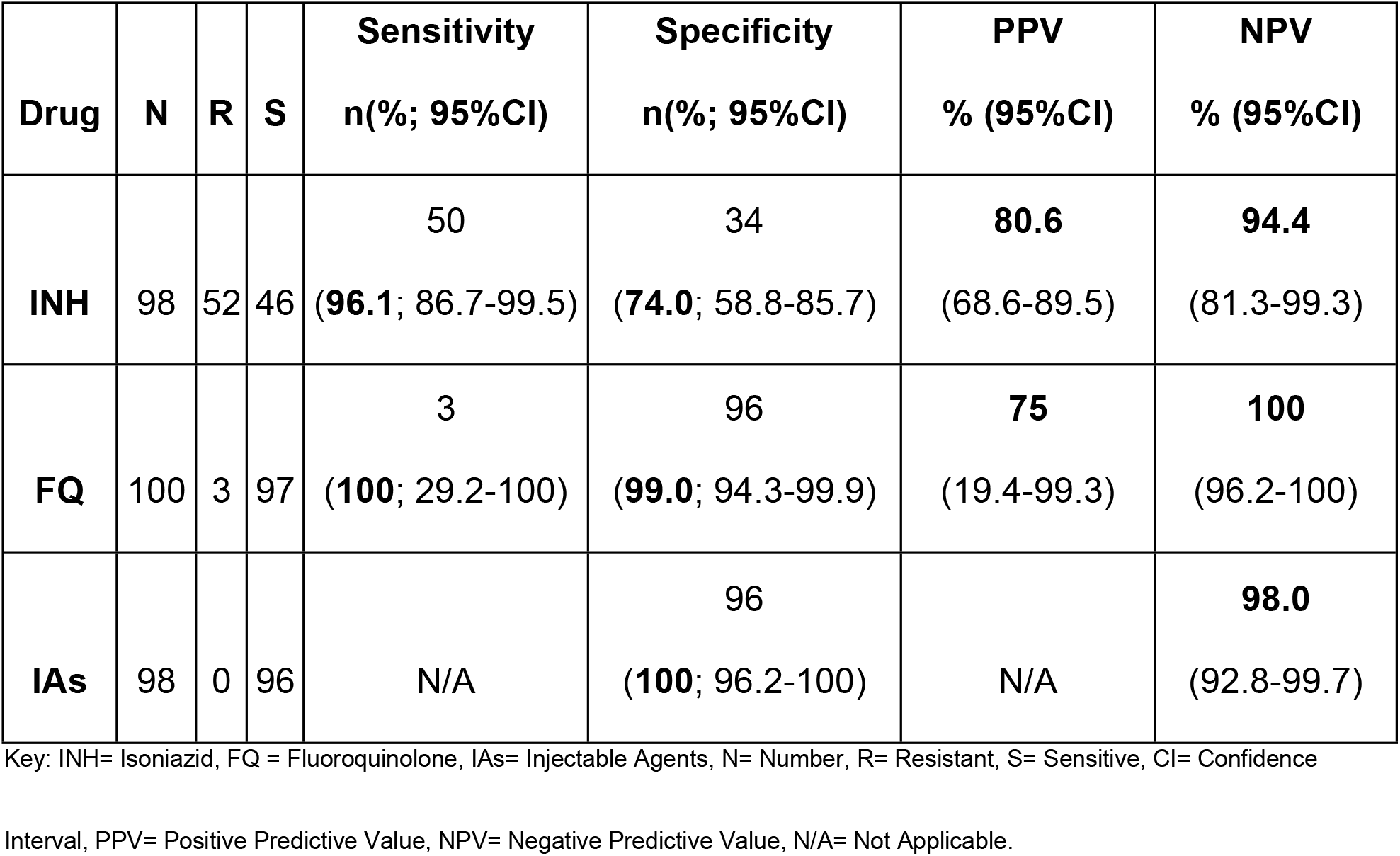
Diagnostic accuracy of Xpert^®^ MTB/XDR test using Hain MDR*plus* and MTBDR*sl* as a reference comparator.

Using LPA as a reference standard, a total of 52/98 (53.1%) were resistant to INH, 3/100 (3.0%) to FQ, and none to IA. The sensitivity and specificity, n (%; 95%CI) of Xpert^®^ MTB/XDR test compared to LPA for; INH was 50 (96.1; 86.7-99.5) and 34 (74.0; 58.8-85.7) and FQ 3 (100; 29.2-100) and 96 (99.0; 94.3-99.9) respectively. All samples were sensitive to AIs leading to, 96 (100; 96.2-100) specificity for AI.

The factors for laboratory uptake were mainly; no training needed for technicians with previous Xpert ultra experience and 1 day for those without, recording and reporting needs were not different from those of Xpert ultra, error rate was 4/100 (4%), no uninterpretable results were recorded, test turn-around-time was 1:45 minutes and workflow similar to that of Xpert ultra.

## Discussion

Results from this in-country validation study show that the Xpert^®^ MTB/XDR test has high sensitivity and specificity regarding isoniazid (INH) and fluoroquinolone (FQs) compared to phenotypic and line probe assay (LPA) drug susceptibility testing (DST). Better diagnostic accuracy measures were recorded when LPA was used as a reference standard. The test achieved acceptable attributes in terms of training needs, test Turn-Around Time (TAT), error rates as well as test workflow. The high sensitivity for both INH and FQs obtained in this study is in agreement with the previous validation studies including a Cochrane review [9-11]. The lower specificity of the Xpert^®^ MTB/XDR test in our study may be attributed to the study design which used the previously recorded results of the reference standard, and in some cases, *M. tuberculosis* isolates were unavailable to repeat the reference test. The End TB strategy’s recommendation for MDR-TB treatment informed by full susceptibility profile remains less implemented in most of the LMICs due to limited accessibility to DST laboratories[4]. The Xpert^®^ MTB/XDR test detects susceptibility to INH, ethionamide, FQs, and IAs (amikacin, kanamycin, and capreomycin), the key drugs used in the management of MDR-TB. It is a semi-quantitative nested PCR followed by high-resolution technology running on the gene Xpert platform. The targets per drug include; INH gene target, *inhA* promotor with -1 to -32 intergenic nucleotide, *katG* codon 311-319, nucleotide 939-957, *fabG1* codon 199-210, nucleotide 597-630, *oxyR-ahpC* with nucleotide -5 to 50 or intergenic -47 to 92. For ethionamide, the gene target is *inhA* promoter with nucleotide -1 to -32 intergenic. For FQs, the gene targets are *gyrA* at codons 87-95 nucleotides 261-285 and *gyrB* at codons 53-544 or 493-505 with nucleotide 1596-1632 [11]. The targeted genes for 2^nd^ line IAs include; amikacin and kanamycin, gene target *rrs* with nucleotide 1396-1417 and capreomycin with gene target *eis* promoter, nucleotides -6 to -42 intergenic [12]. The Xpert MTB/XDR test offers DST results in less than 90 minutes with limited technical and infrastructural requirements. The test exhibits better technical and operational attributes compared to other WHO-endorsed molecular diagnostics for DR-TB [13, 14]. This makes the test ideal for reducing the current challenges of obtaining INH and FQs DST results for better DR-TB patient management. The WHO recommends that in people with bacteriologically confirmed pulmonary TB, low complexity automated NAATs may be used on sputum for the initial detection of resistance to INH and FQ, rather than culture-based phenotypic DST. Early diagnosis and DST forms a stronger pillar for DR-TB management and control [4]. Molecular rapid diagnosis/DST followed by rapid appropriate treatment initiation provides greater benefits for patient management as well as DR-TB control. The TB diagnostic algorithm including Xpert MTB/XDR test will reduce the diagnostic challenges needed for tests that are requested from the reference laboratories such as LPA and culture and DST[2]. In most of the LMICs, TB patients are initiated on treatment with limited or no DST results. If results of FQ DTS are not available at the initiation of DR-TB treatment, it may increase the risk of acquired bedaquiline resistance in undetected FQ resistance mainly in newly endorsed BPaLM regimen[15]. In some areas over 30% of the RR-TB patients are already resistant to FQ, pre-XDR, at the start of the treatment[16]. Therefore, implementing the Xpert MTB/XDR test means that more patients will be initiated to DST guided TB treatment. This is likely to lead to better MDR-TB treatment outcomes as well as better estimates of resistance to INH and FQs the two key drugs in the current standard regimens for the management of DR-TB[2].

Our study had some limitations: First the number of samples with resistance to FQs and injectable agents were low given the fact that Uganda is a low MDR/RR-TB setting, however, this increases confidence in specificity and makes it a suitable setting where Xpert MTB/XDR test is most needed. We used previously stored sputum and/or isolates which may have reduced the sensitivity, however, the direct testing was done on molecular tests and indirect on cultured isolates. Some of the data was missing and we could not relate the accuracy to patient’s clinical and demographic data. Our study did not test for ethionamide, a drug also included in the Xpert MTB/XDR test and in the management of MDR/RR-TB, because this is not tested phenotypically in Uganda, however, a recent study found DST for ethionamide to have suboptimal sensitivity on Xpert MTB/XDR test due to inclusion of only mutations in the *inhA* promoter region[17].

## Conclusion

The Xpert MTB/XDR test is a reliable, rapid accurate, and easy-to-use DST method for isoniazid, fluoroquinolone, and injectable agents. We recommend a first roll-out phase to consider prioritization of all DR-TB treatments to have 10-color machines which can run both Xpert Ultra and XDR tests. For better uptake and roll-out, countries should consider all new GeneXpert machine procurements to be 10-color for easy integration. In the future, the manufacturer should plan to swap 6-color with 10-color Xpert modules in the existing GeneXpert Ultra machines to ensure better access to XDR test in all Xpert sites.

## Data Availability

All relevant data are within the manuscript and its Supporting Information files.

## Supporting information

**S1File:** Dataset for the validation of Xpert MTB/XDR test in Uganda

## Data availability statement

All relevant data are within the paper and its supporting information files

## Acknowledgment

We thank the Mycobacteriology (BSL-3) Laboratory at Makerere University and the Supranational TB reference Laboratory (SRL) Uganda for the sputum and isolates provided for this study. We acknowledge the support from Cepheid for technical support, the Xpert MTB/XDR machine and cartridges used in this validation study. Cepheid had to influence in the design and conduct of this validation study in Uganda. WS is a NURTURE fellow under NIH grant D43TW010132, a postdoctoral fellow under MUII+, Uganda Medical Informatics Centre (UMIC) Bioinformatics endeavour and under the European & Developing Countries Clinical Trials Partnership (EDCTP) grant TMA2018CDF-2351. The funders had no role in study design, data collection and analysis, decision to publish, or preparation of the manuscript.

## Author Contributions

**Conceptualization:** Achilles Katamba, Willy Ssengooba and Moses Joloba

**Data curation:** Achilles Katamba, Willy Ssengooba, James Serubiri, Derrick Semugenze, Kasule George William, Nyombi Abdunoor, Raymond Byaruhanga, Stavia Turyahabwe and Moses L Joloba

**Formal Analysis:** Achilles Katamba, Willy Ssengooba, and Moses L Joloba

**Funding Acquisition:** Achilles Katamba, Willy Ssengooba, Moses L Joloba, Byaruhanga, Stavia Turyahabwe

**Investigation:** Achilles Katamba, Willy Ssengooba, James Serubiri, Derrick Semugenze, Kasule George William, Nyombi Abdunoor, Raymond Byaruhanga, Stavia Turyahabwe and Moses L Joloba

**Methodology:** Achilles Katamba, Willy Ssengooba, James Serubiri, Derrick Semugenze, Kasule George William, Nyombi Abdunoor, Raymond Byaruhanga, Stavia Turyahabwe and Moses L Joloba

**Project administration:** Achilles Katamba, Willy Ssengooba, and Moses L Joloba **Resources:** Achilles Katamba, Willy Ssengooba, and Moses L Joloba **Supervision:** Achilles Katamba, Willy Ssengooba, and Moses L Joloba

**Validation:** Achilles Katamba, Willy Ssengooba, Moses L Joloba Byaruhanga, Stavia Turyahabwe

**Visualization:** Achilles Katamba, Willy Ssengooba, Moses L Joloba

**Writing original draft:** Achilles Katamba, Willy Ssengooba, Moses L Joloba

**Writing, Review and editing:** Achilles Katamba, Willy Ssengooba, James Serubiri, Derrick Semugenze, Kasule George William, Nyombi Abdunoor, Raymond Byaruhanga, Stavia Turyahabwe and Moses L Joloba

## Conflict of interest

The authors have declared that no competing interests exist

## Notes

### Competing Interest Statement

The authors have declared no competing interest.

### Author Declarations

The study received ethics committee approval from the Makerere University School of Biomedical Sciences Research Ethics Committee and the Uganda National Council of Science and Technology.

